# Violation of the constant genetic effect assumption can result in biased estimates for non-linear Mendelian randomization

**DOI:** 10.1101/2022.10.26.22280570

**Authors:** Stephen Burgess

## Abstract

Non-linear Mendelian randomization is an extension of conventional Mendelian randomization that performs separate instrumental variable analyses in strata of the study population with different average levels of the exposure. The approach estimates a localized average causal effect function, representing the average causal effect of the exposure on the outcome at different levels of the exposure. The commonly-used residual method for dividing the population into strata works under the assumption that the effect of the genetic instrument on the exposure is linear and constant in the study population. However, this assumption may not hold in practice. We use the recently developed doubly-ranked method to re-analyse various datasets previously analysed using the residual method. In particular, we consider a genetic score for 25-hydroxyvitamin D [25(OH)D] used in a recent non-linear Mendelian randomization analysis to assess the potential effect of vitamin D supplementation on all-cause mortality. We show that the effect of the genetic score on 25(OH)D concentrations varies strongly, with a five-fold difference in the estimated genetic association with the exposure in the lowest and highest decile groups. Evidence for a protective causal effect of vitamin D supplementation on all-cause mortality in low vitamin D individuals is evident for the residual method, but not for the doubly-ranked method. We show that the constant genetic effect assumption is more reasonable for some exposures, and less reasonable for others. If the doubly-ranked method indicates that this assumption is violated, then estimates from both the residual and doubly-ranked methods can be biased, although bias was smaller on average in the doubly-ranked method. Analysts should compare results from both methods, as well as considering transforming the exposure to reduce heterogeneity in the genetic effect on the exposure.

## Introduction

Mendelian randomization is an epidemiological approach that uses genetic variants as instrumental variables to assess the potential causal effect of an exposure on an outcome [1]. Conventional instrumental variable analyses target either an average treatment effect or a local average treatment effect [2]. Non-linear Mendelian randomization is an extension to conventional Mendelian randomization for use with a continuous exposure that conducts separate instrumental variable analyses in strata of the study population with different average values of the exposure [3]. The motivation is to investigate the shape of the localized average causal effect curve for the exposure on the outcome at different values of the exposure [4]

Stratification on the exposure directly would lead to collider bias, as the exposure is a collider in the standard instrument variable directed acyclic graph (Figure 1); it is a common effect of the genetic variants and confounders [5]. Two methods have been proposed to stratify the population while avoiding collider bias: a residual method [3] and a doubly-ranked method [6]. The residual method is implemented by first regressing the exposure on the genetic instrument, and taking the residual from this regression. This ‘residual exposure’ is now independent of the genetic instrument, and so strata based on the residual exposure are uncorrelated with the instrument [7]. The doubly-ranked method is implemented by first ranking participants into pre-strata according to their level of the genetic instrument, and then by ranking participants within each pre-stratum according to their level of the exposure. For example, if we want to divide the population into 10 strata, then each pre-stratum should contain 10 individuals. Then, the individual with the lowest level of the exposure from each pre-stratum is selected into stratum 1, the individual with the next lowest level of the exposure from each pre-stratum is selected into stratum 2, and so on, so that each stratum includes one individual from each pre-stratum [6].

**Figure 1:**
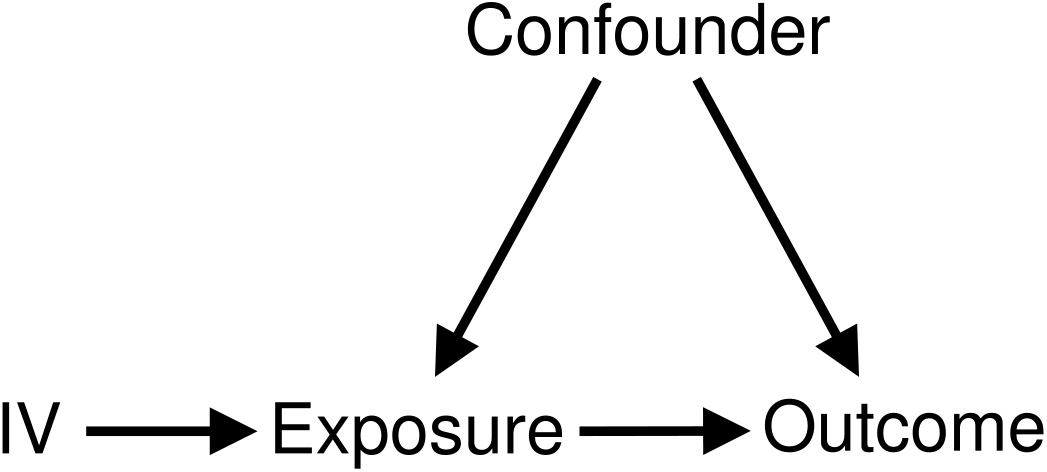
Directed acyclic graph of instrumental variable (IV) assumptions

While the residual method was developed first and has generally been used in applied practice [8, 9, 10, 11, 12, 13, 14, 15], it has some limitations. In particular, it assumes that the effect of the genetic instrument on the exposure is linear and constant for all individuals in the population. This is a strong assumption [4]. Although the initial exposition of this method did investigate sensitivity to this assumption [3], the investigation was limited in scope – only moderate violations of this assumption were considered. Subsequent investigations have shown that variability in the genetic effect can lead to bias and inflation of Type 1 error rates [6]. In contrast, the doubly-ranked method does not make any parametric assumption on the effect of the genetic instrument. A weakness of the doubly-ranked method is that the strata are not defined with respect to clinically relevant cut-offs of the exposure distribution, and so the interpretation of stratum-specific estimates is less clear.

In this manuscript, we revisit a previously published analysis into the effect of vitamin D supplementation on all-cause mortality risk that was undertaken by non-linear Mendelian randomization using the residual method [16]. We show that the assumption of a constant genetic effect on the exposure does not appear to be satisfied in this case. When re-analysing the data using the doubly-ranked method, evidence for a protective effect of vitamin D supplementation on all-cause mortality in low vitamin D individuals is substantially attenuated, and the estimate for this stratum is compatible with no effect. A similar finding is observed from the residual method when log-transforming the exposure; log-transformation could reduce the difference between the magnitude of the genetic effect on the exposure at different exposure levels. Although the genetic associations with the exposure often seem homogeneous within strata defined by the residual method, this similarity appears to be artefactual rather than a true guide of the validity of the constant genetic effect assumption. We conclude by discussing the interpretation of findings from non-linear Mendelian randomization in the light of this analysis.

## Methods

### Simulation study 1

To demonstrate the potential for bias when the constant genetic effect assumption is violated, we conduct an illustrative simulation study. We generate data on a binary genetic instrument *G*, a binary confounder *U*, and a continuous exposure *X*. We compare the distribution of the confounder within strata of the exposure for different values of the genetic instrument. In conventional Mendelian randomization, the instrument should be independent of the confounder in the study population. To avoid bias in non-linear Mendelian randomization, the distribution of the confounder should be the same in the *G* = 0 and *G* = 1 groups of the population for each stratum. The distribution of the confounder will vary between strata, but for a given stratum, the proportion of individuals with *U* = 0 and *U* = 1 should be the same for the genetic group with *G* = 0 as with *G* = 1.

We generate data according to the following model for 1 000 000 individuals indexed by *i*:

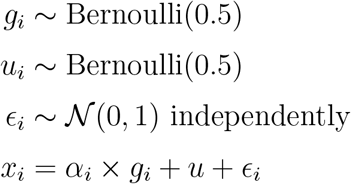

The genetic instrument *G* takes value 0 or 1 at random with probability 0.5. The confounder *U* independently takes value 0 or 1 at random with probability 0.5. The exposure is a linear combination of the genetic instrument, confounder, and an independent error term. We consider three scenarios for the genetic effect on the exposure *α*_*i*_:

1. **Constant genetic effect:** *α*_*i*_ = 0.4. The genetic effect is 0.4 for all individuals in the population.
2. **Independent varying genetic effect:** *α*_*i*_ ∼ 𝒩 (0.4, 0.2^2^). For each individual, the genetic effect is drawn from a normal distribution with mean 0.4 and standard deviation 0.2, independently of the confounder.
3. **Dependent varying genetic effect:** *α*_*i*_ ∼𝒩 (0.2, 0.1^2^) if *u*_*i*_ = 0, and *α*_*i*_ ∼𝒩 (0.6, 0.1^2^) if *u*_*i*_ = 1. For individuals with *U* = 0, the genetic effect is drawn from a normal distribution with mean 0.2 and standard deviation 0.1, and for individuals with *U* = 1, the genetic effect is drawn from a normal distribution with mean 0.6 and standard deviation 0.1. Therefore the effect of the genetic variant depends on the value of the confounder.

In each scenario, we consider the distribution of the confounder in 10 strata formed by stratifying directly on the exposure versus those formed by stratifying on the residual exposure.

### Simulation study 2

To explore the direction of bias when the genetic effect on the exposure varies, we consider a further simulation study. In this case, to compare results from the residual and doubly-robust methods, we simulate data on a continuous genetic instrument *G* and a continuous confounder *U*, each drawn from independent standard normal distributions. For each stratum, we compare the proportion of individuals with *U* < 0 and *U* > 0 for the genetic group with *G* < 0 versus *G* > 0.

We generate data according to the following model for 1 000 000 individuals indexed by *i*:

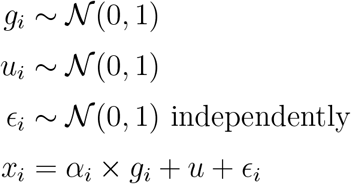

We consider three scenarios:

1. **Genetic effect depends on** *X* **only:** *α*_*i*_ ∼ *N* (0.3 + 0.1 × *ϵ*_*i*_, 0.1^2^). The genetic effect depends on the value of the error term *ϵ*, and hence only on the component of *X* that is independent of *U*.
2. **Genetic effect depends of** *X* **and** *U* **equally:**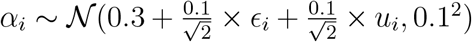. The genetic effect depends equally on the values of the error term *ϵ* and the confounder *U*.
3. **Genetic effect depends on** *U* **only:** *α*_*i*_ ∼ *N* (0.3 + 0.1 × *u*_*i*_, 0.1^2^). The genetic effect depends on the value of the confounder *U*.

In each scenario, we consider the distribution of the confounder in 10 strata formed by stratifying on the residual exposure versus those formed by the doubly-ranked method. We also conduct a more systematic simulation study to investigate the degree of potential bias in the residual versus doubly-ranked method; details are provided in the Supplementary Material.

### Re-analysis of UK Biobank data

We follow the genetic analyses previously presented in the Sofianopoulou paper [16] using data from UK Biobank on 333 002 individuals. The exposure is concentration of 25-hydroxyvitamin D [25(OH)D], a circulating marker of vitamin D status, corrected for season of blood draw. The genetic instrument is a score comprising 21 genetic variants from 4 gene regions related to vitamin D synthesis and metabolism. The outcome is all-cause mortality. More detailed definitions and descriptions of the analytic sample can be found in the original paper [16]. We perform the residual and doubly-ranked methods, dividing the population into 10 equal-sized strata, and estimate genetic associations with the exposure and outcome within each stratum using linear regression for the exposure, and logistic regression for the outcome, with adjustment for age, sex, centre, and 10 genomic principal components.

We also conduct analyses on log-transformed values of the exposure. Log-transformation will not change the order of participants when ranked by the exposure, and so associations with the outcome from the doubly-ranked method will be unchanged. However, it will affect the constant genetic effect assumption in the residual method, as the assumption is now that the genetic effect is constant on a log-scale. This could be plausible if the genetic variants affect the exposure in a proportional way. In any case, if the genetic effects are larger on the untransformed scale for individuals with greater concentrations of 25(OH)D, a log-transformation will reduce this heterogeneity.

To explore the validity of the stratification, we assess the association between the genetic instrument and a small number of covariates that are potential confounders: low-density lipoprotein (LDL) cholesterol, glycated haemoglobin (HbA1c), and forced expiratory volume in the first second (FEV1) in strata defined by the residual and doubly-ranked methods.

Finally, to investigate the plausibility of the constant genetic effect assumption for different exposures, we re-analysed UK Biobank data that have previously been analysed by non-linear Mendelian randomization using the residual method, but instead using the doubly-ranked method. We considered as exposures glycated haemoglobin (HbA1c) [8], systolic blood pressure (SBP) [9], body mass index (BMI) [10], and kidney function (presented as estimated glomerular filtration rate, eGFR) using the same genetic instruments as presented in the original studies.

## Results

### Simulation study 1

Results from the first simulation study are shown in Figure 2: the top panel shows results stratifying on the exposure directly, whereas the bottom panel shows results stratifying on the residual exposure. For each stratum, the proportion of individuals with *U* = 0 is displayed in red, and the proportion of individuals with *U* = 1 is displayed in blue. If the stratification is valid, the proportion of individuals with *U* = 0 should be the same in both groups of the genetic instrument (light red for *G* = 0, dark red for *G* = 1); similarly for *U* = 1 (light blue for *G* = 0, dark blue for *G* = 1). We note that *G* and *U* were generated as independent random variables, and so their distributions will be independent in the population as a whole.

**Figure 2:**
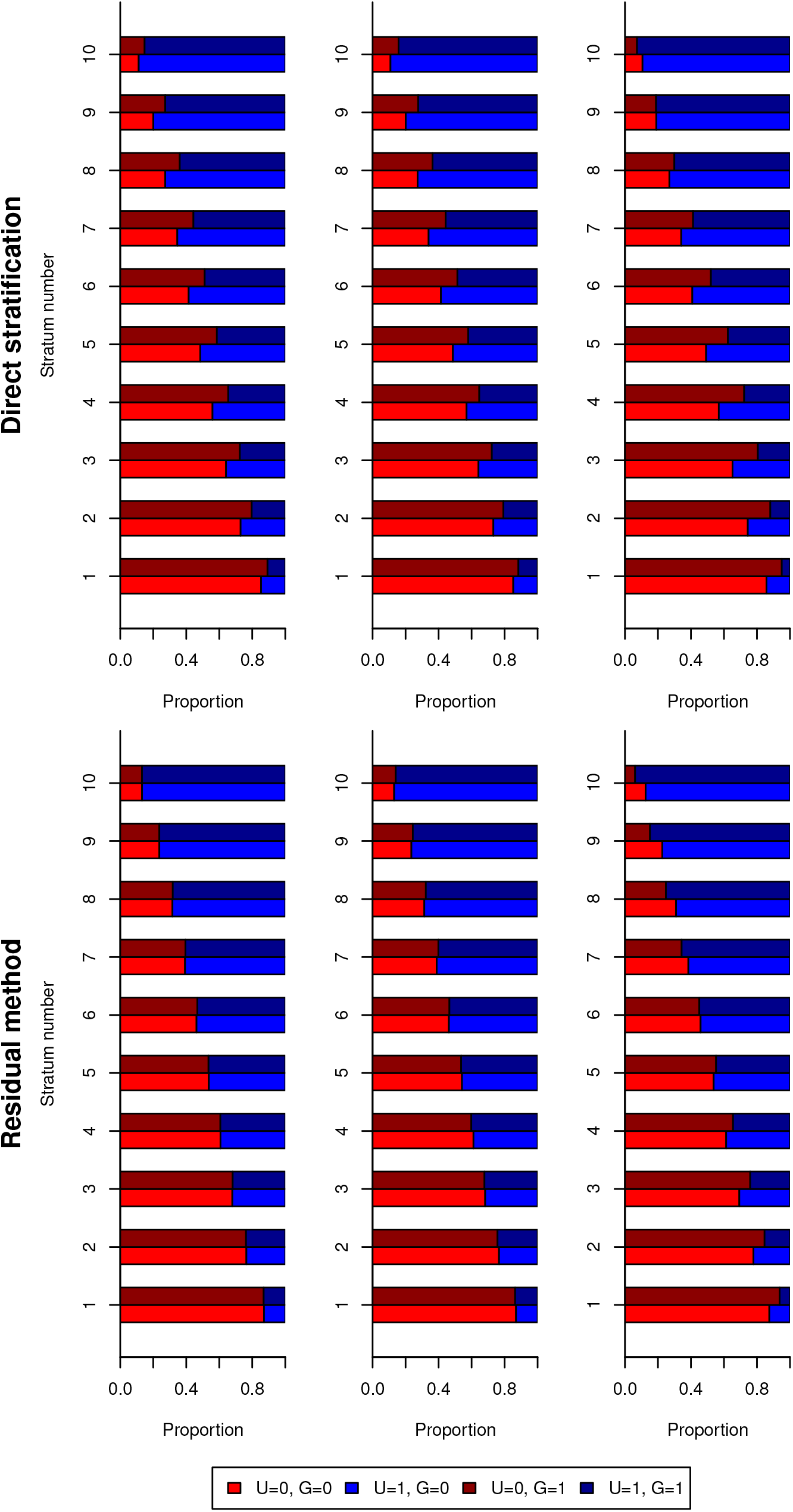
Simulation study 1 results, indicating the proportion of individuals in each genetic group (*G* = 0, brighter colours; or *G* = 1, darker colours) for each stratum with the confounder absent (*U* = 0, red) or present (*U* = 1, blue). Strata are defined by stratification on the exposure directly (top panel) or on the residual exposure (bottom panel). For each stratification method, results are presented in three scenarios (left-to-right): 1) constant genetic effect, 2) independent varying genetic effect, and 3) dependent varying genetic effect.

In the top panel, we see the distribution of the confounder *U* differs between genetic groups in each stratum when stratifying on the exposure directly. This difference is evident in all three simulation scenarios, although it is strongest in Scenario 3 (dependent varying genetic effect). This is due to collider bias; once we stratify on the exposure, the instrument and confounder are dependent within strata conditional on the exposure. In the bottom panel, when stratifying on the residual exposure the distribution of the confounder is the same for both genetic groups in scenarios 1 (constant genetic effect) and 2 (independent varying genetic effect). However, the distribution of the confounder differs between genetic groups in Scenario 3. In Scenario 1, the residual exposure is no longer a function of the genetic instrument, and hence it is no longer a collider, and the instrument and confounder remain independent within strata conditional on the residual exposure. But if the effect of the genetic instrument on the exposure is not constant, then the functional relationship between the instrument and the exposure is not fully broken, and so the residual exposure is still a collider. This simulation study indicates that there will typically be bias in non-linear Mendelian randomization estimates from the residual method when the effect of the genetic instrument on the exposure varies, particularly when this variability depends on the value of a confounder.

### Simulation study 2

Results from the second simulation study are shown in Figure 3: the top-left panel shows results stratifying on the exposure directly, the top-right panel shows results stratifying on the residual exposure, and the bottom panel shows results from the doubly-ranked stratification. Similarly to the previous figure, the proportion of individuals with *U* < 0 is displayed in red, and the proportion of individuals with *U* > 0 is displayed in blue. We would like the proportion of individuals with *U* < 0 to be the same in both groups of the genetic instrument (light red for *G* < 0, dark red for *G* > 0); similarly for *U* > 0 (light blue for *G* < 0, dark blue for *G* > 0).

**Figure 3:**
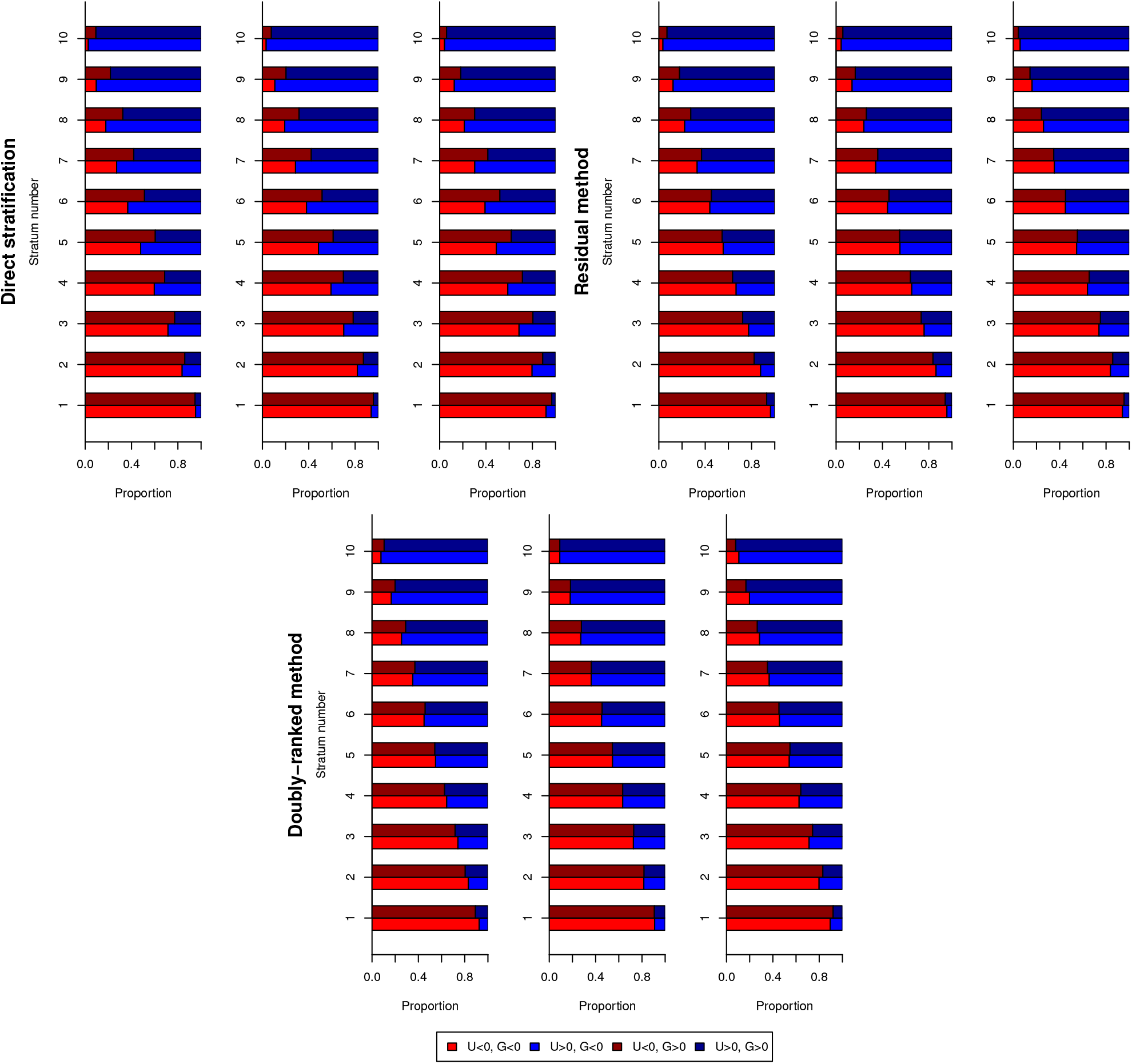
Simulation study 2 results, indicating the proportion of individuals in each genetic group (*G* = 0, brighter colours; or *G* > 0, darker colours) for each stratum with the confounder below (*U* < 0, red) or above (*U* > 0, blue) its median value. Strata are defined by stratification on the exposure directly (top-left panel), the residual exposure (top-right panel) or using the doubly-ranked method (bottom panel). For each stratification method, results are presented in three scenarios (left-to-right): 1) genetic effect depends on *X* only, 2) genetic effect depends on *X* and *U*, and 3) genetic effect depends on *U* only.

Again, when stratifying on the exposure directly, there is a substantial difference in the distribution of the confounder between the genetic groups for each stratum. This difference is reduced in the residual method, and reduced further in the doubly-ranked method. However, even in the doubly-ranked method, some difference remains in Scenarios 1 and 3. We note that the direction of the difference varies between Scenarios 1 and 3 for both the residual and doubly-ranked methods; in Scenario 1, the instrument is positively correlated with the confounder in low strata and negatively correlated in high strata, whereas in Scenario 3, the instrument is negatively correlated with the confounder in low strata and positively correlated in high strata. This indicates that violation of the constant genetic effect assumption could bias Mendelian randomization estimates in either the positive or negative direction.

We also conducted a more systematic investigation to explore the correlation between the confounder and genetic instrument in strata from the residual and doubly-ranked methods in a range of scenarios (Supplementary Material). In each scenario, for both methods the average degree of correlation depended on the variability of the genetic effects on the exposure but was independent of average instrument strength, and the correlation was on average two times larger for the residual method compared with the doubly-ranked method (Supplementary Table A1).

### Vitamin D and all-cause mortality

Genetic associations from the residual and doubly-ranked methods are presented in Table 1. While genetic associations with the exposure are similar in strata defined by the residual method (with the exception of the highest and lowest strata), the estimates are suspiciously similar with a consistent evident trend. Simulations have shown that these association estimates can be similar even when the true genetic effects vary [6]. In contrast, genetic associations with the exposure in strata defined by the doubly-ranked method differ strongly, with a five-fold difference between estimates in the lowest versus highest stratum. This suggests that the constant genetic effect assumption is questionable in this example.

**Table 1:** Genetic associations in strata of the population defined by the residual and doubly-ranked methods for the exposure of 25(OH)D: for each stratum, we present the mean 25(OH)D concentration (nmol/L; in brackets, 10th and 90th percentiles of the distribution, or 20th and 80th percentiles for the lowest and highest strata); the association of the genetic instrument with the exposure (nmol/L; in brackets, its standard error); and the association of the genetic instrument with the outcome (log odds ratio; in brackets, its standard error).

| Residual method |  |  |  |
| --- | --- | --- | --- |
| Stratum | 25(OH)D concentration | Association with exposure | Association with outcome |
| 1 | 26.8 (22.2, 33.9) | 15.02 (0.14) | −0.508 (0.094) |
| 2 | 34.9 (28.8, 40.8) | 18.68 (0.05) | −0.350 (0.104) |
| 3 | 40.1 (33.8, 46.2) | 18.99 (0.04) | −0.084 (0.105) |
| 4 | 45.0 (38.5, 51.1) | 19.19 (0.03) | −0.014 (0.106) |
| 5 | 49.8 (43.2, 56.0) | 19.16 (0.03) | 0.031 (0.107) |
| 6 | 54.7 (48.0, 61.0) | 19.25 (0.03) | −0.148 (0.106) |
| 7 | 60.2 (53.5, 66.3) | 19.25 (0.04) | 0.027 (0.111) |
| 8 | 66.4 (59.7, 72.7) | 19.34 (0.05) | −0.059 (0.115) |
| 9 | 74.6 (67.5, 81.3) | 19.55 (0.07) | −0.231 (0.121) |
| 10 | 92.1 (79.5, 100.2) | 24.08 (0.30) | 0.053 (0.126) |

Table 1: Genetic associations in strata of the population defined by the residual and doubly-ranked methods for the exposure of 25(OH)D: for each stratum, we present the mean 25(OH)D concentration (nmol/L; in brackets, 10th and 90th percentiles of the distribution, or 20th and 80th percentiles for ...
| Doubly-ranked method |  |  |  |
| --- | --- | --- | --- |
| Stratum | 25(OH)D concentration | Association with exposure | Association with outcome |
| 1 | 28.6 (22.6, 38.1) | 6.82 (0.18) | −0.020 (0.089) |
| 2 | 35.7 (27.0, 45.0) | 9.22 (0.17) | 0.079 (0.100) |
| 3 | 41.1 (32.0, 51.0) | 11.70 (0.18) | 0.068 (0.105) |
| 4 | 45.9 (36.1, 56.5) | 14.38 (0.18) | 0.145 (0.110) |
| 5 | 50.7 (40.2, 61.9) | 17.00 (0.19) | −0.150 (0.110) |
| 6 | 55.5 (44.3, 67.4) | 19.78 (0.20) | 0.109 (0.111) |
| 7 | 60.6 (48.6, 73.3) | 22.64 (0.21) | −0.110 (0.113) |
| 8 | 66.5 (53.4, 80.1) | 25.86 (0.22) | −0.141 (0.112) |
| 9 | 73.8 (59.0, 89.4) | 29.29 (0.25) | −0.009 (0.114) |
| 10 | 86.2 (67.2, 98.6) | 34.14 (0.36) | −0.003 (0.112) |

Although there is a strong association between the genetic score and the outcome in the lowest stratum defined by the residual method (*β* = *−*0.508, p-value 6*×*10^*−*8^), no association is evident in the lowest stratum defined by the doubly-ranked method (*β* = *−*0.020, p-value 0.82). These estimates can be converted to an odds ratio (OR) per 10 nmol/L higher genetically-predicted 25(OH)D of 0.71 (95% confidence interval [CI] 0.63, 0.81) for the residual method, and 0.97 (95% CI 0.75, 1.25) for the doubly-ranked method. There was also an association with the outcome in the second lowest stratum (stratum 2) defined by the residual method, although slightly weaker in magnitude (*β* = *−*0.350, p-value 0.0007; OR 0.83, 95% CI 0.74, 0.92); again, this inverse association was not observed in the doubly-ranked method. The confidence intervals for the doubly-ranked method are wider in lower strata and narrower in higher strata; this is consistent with the genetic effect on the exposure being stronger in higher strata.

Genetic associations when the exposure is log-transformed are presented in Table 2. As expected, genetic associations with the outcome in strata defined by the doubly-ranked method are unaffected, as log-transformation does not change the ranking of participants. Genetic associations with the exposure still vary, but the difference is far less pronounced, with a less than two-fold difference (1.72-fold) between estimates in the lowest stratum versus the highest stratum for the doubly-ranked method. For the residual method, genetic associations with the exposure retain the same pattern, but the genetic association with the outcome in the lowest stratum was substantially attenuated to the null (*β* = *−*0.156, p-value 0.064). This represents an odds ratio of 0.86 (95% CI 0.73, 1.01) per 40% increase in genetically-predicted 25(OH)D. A 40% increase represents the increase from 25 nmol/L to 35 nmol/L, and so is comparable to a 10 nmol/L increase for the lowest stratum.

**Table 2:** Genetic associations in strata of the population defined by the residual and doubly-ranked methods for the exposure of log-transformed 25(OH)D: for each stratum, we present the mean 25(OH)D concentration (log-transformed nmol/L; in brackets, 10th and 90th percentiles of the distribution, or 20th and 80th percentiles for the lowest and highest strata); the association of the genetic instrument with the exposure (log-transformed nmol/L; in brackets, its standard error); and the association of the genetic instrument with the outcome (log odds ratio; in brackets, its standard error).

| Residual method |  |  |  |
| --- | --- | --- | --- |
| Stratum | Log-transformed 25(OH)D | Association with exposure | Association with outcome |
| 1 | 3.23 (3.09, 3.46) | 0.337 (0.006) | -0.156 (0.084) |
| 2 | 3.54 (3.41, 3.67) | 0.334 (0.001) | 0.033 (0.099) |
| 3 | 3.69 (3.57, 3.81) | 0.339 (0.001) | -0.054 (0.105) |
| 4 | 3.81 (3.69, 3.92) | 0.340 (0.001) | 0.166 (0.108) |
| 5 | 3.91 (3.79, 4.02) | 0.340 (0.001) | -0.041 (0.107) |
| 6 | 4.01 (3.89, 4.12) | 0.342 (0.001) | -0.056 (0.109) |
| 7 | 4.10 (3.98, 4.21) | 0.342 (0.001) | -0.052 (0.113) |
| 8 | 4.20 (4.08, 4.31) | 0.343 (0.001) | -0.033 (0.116) |
| 9 | 4.31 (4.19, 4.42) | 0.343 (0.001) | -0.151 (0.122) |
| 10 | 4.50 (4.34, 4.61) | 0.345 (0.003) | -0.026 (0.124) |

Table 2: Genetic associations in strata of the population defined by the residual and doubly-ranked methods for the exposure of log-transformed 25(OH)D: for each stratum, we present the mean 25(OH)D concentration (log-transformed nmol/L; in brackets, 10th and 90th percentiles of the distribution, ...
| Doubly-ranked method |  |  |  |
| --- | --- | --- | --- |
| Stratum | Log-transformed 25(OH)D | Association with exposure | Association with outcome |
| 1 | 3.31 (3.12, 3.64) | 0.240 (0.007) | -0.020 (0.089) |
| 2 | 3.55 (3.30, 3.81) | 0.258 (0.005) | 0.079 (0.100) |
| 3 | 3.70 (3.46, 3.93) | 0.284 (0.004) | 0.068 (0.105) |
| 4 | 3.81 (3.59, 4.03) | 0.314 (0.004) | 0.145 (0.110) |
| 5 | 3.91 (3.69, 4.13) | 0.338 (0.004) | -0.150 (0.110) |
| 6 | 4.00 (3.79, 4.21) | 0.361 (0.004) | 0.109 (0.111) |
| 7 | 4.09 (3.88, 4.30) | 0.380 (0.003) | -0.110 (0.113) |
| 8 | 4.18 (3.98, 4.38) | 0.398 (0.003) | -0.141 (0.112) |
| 9 | 4.29 (4.08, 4.49) | 0.408 (0.003) | -0.009 (0.114) |
| 10 | 4.44 (4.21, 4.59) | 0.412 (0.004) | -0.003 (0.112) |

Genetic associations with the covariates in strata of the population are shown in Figure 4. In the overall population, associations with all three traits are null: LDL-cholesterol, *β* = *−*0.000, *p* = 0.96; HbA1c, *β* = 0.011, *p* = 0.16; FEV1, *β* = 0.003, *p* = 0.59. Beta-coefficients represent the association with the covariate in standard deviation units per unit change in the genetic instrument, corresponding to a standard deviation change in 25(OH)D concentrations. However, there are evident associations with covariates within several strata of the population defined by the residual method: for LDL-cholesterol, there are strong associations in higher strata, and for HbA1c and FEV1, there are strong associations in lower strata. In contrast, genetic associations with covariates are less strong in strata defined by the doubly-ranked method. When log-transforming 25(OH)D, associations with covariates in strata from the residual method were attenuated (Supplementary Figure A1). This provides a further line of evidence questioning results from the residual method for untransformed values of 25(OH)D concentrations.

**Figure 4:**
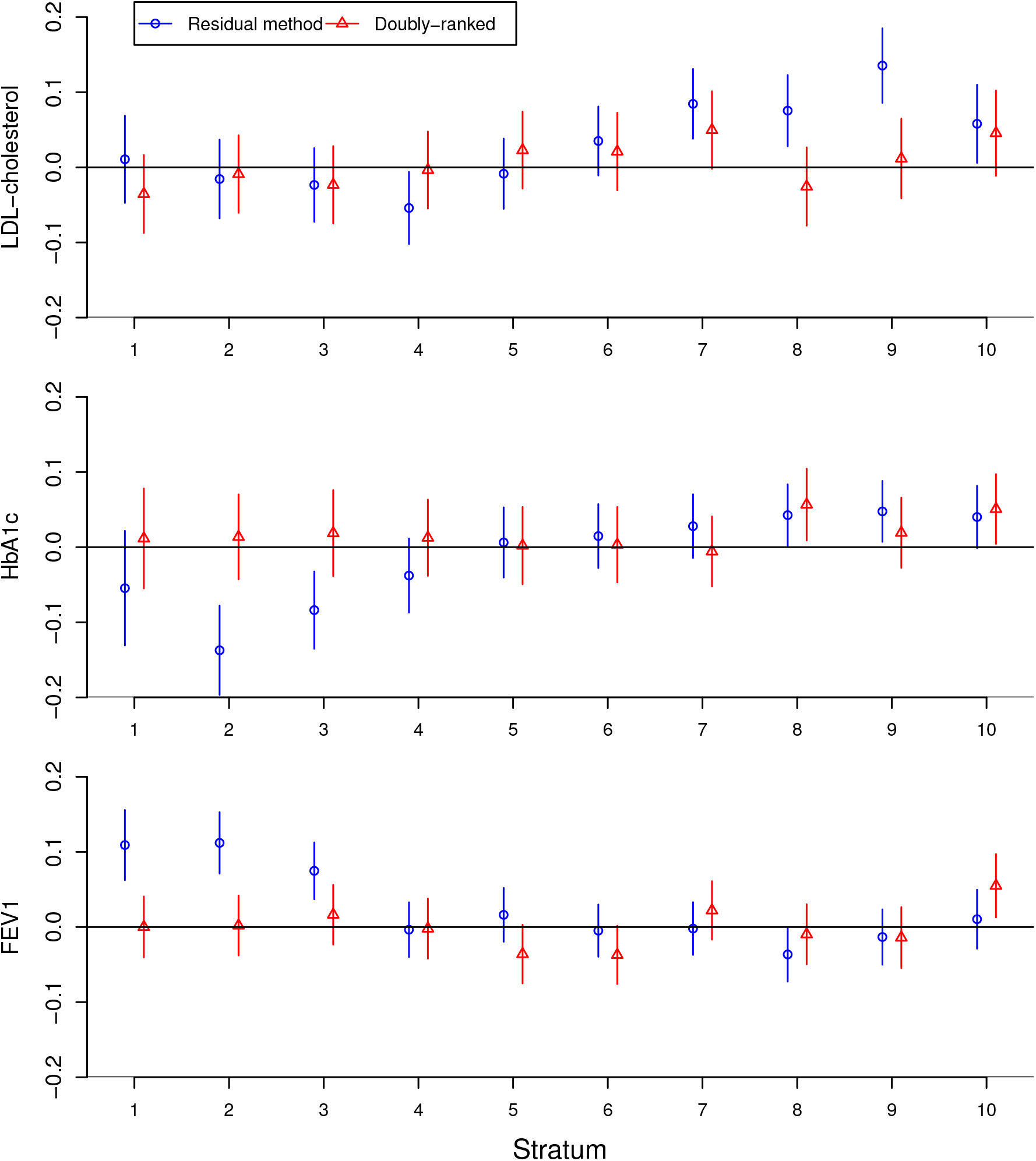
Genetic associations with covariates in strata defined by the residual method (blue circles) and doubly-ranked method (red triangles). Estimates represent associations with the covariate in standard deviation units per unit change in the genetic instrument, corresponding to a standard deviation change in 25(OH)D concentrations. Error bars are 95% confidence intervals.

### Other exposures

To assess the plausibility of the constant genetic effect assumption more broadly, we performed similar analyses using the doubly-ranked method for other exposures. Genetic associations with the exposure in strata defined by the doubly-ranked method are given in Table 3 for HbA1c, SBP, BMI, and eGFR.

**Table 3:** Genetic associations in strata of the population defined by the doubly-ranked method for the exposures of glycated haemoglobin (HbA1c), systolic blood pressure (SBP), body mass index (BMI), and estimated glomerular filtration rate (eGFR). Genetic associations with the exposure (in brackets, standard errors) are expressed in mmol/mol for HbA1c, mmHg for SBP, kg/m^2^ for BMI, and mL/min/1.73m^2^ for eGFR.

| Stratum | Association with ... |  |  |  |
| --- | --- | --- | --- | --- |
|  | HbA1c | SBP | BMI | eGFR |
| 1 | 10.93 (0.53) | 0.679 (0.015) | 2.03 (0.06) | 2.749 (0.055) |
| 2 | 10.28 (0.35) | 0.763 (0.013) | 2.43 (0.05) | 2.908 (0.038) |
| 3 | 10.65 (0.30) | 0.815 (0.012) | 2.77 (0.05) | 2.861 (0.033) |
| 4 | 11.18 (0.28) | 0.863 (0.012) | 3.17 (0.05) | 2.678 (0.029) |
| 5 | 11.17 (0.27) | 0.906 (0.013) | 3.45 (0.05) | 2.402 (0.026) |
| 6 | 11.60 (0.27) | 0.926 (0.013) | 3.73 (0.06) | 2.126 (0.023) |
| 7 | 11.77 (0.28) | 0.952 (0.014) | 4.12 (0.07) | 1.910 (0.023) |
| 8 | 11.96 (0.30) | 0.970 (0.016) | 4.78 (0.08) | 1.719 (0.023) |
| 9 | 12.71 (0.35) | 0.994 (0.019) | 5.62 (0.10) | 1.516 (0.023) |
| 10 | 13.24 (0.48) | 1.059 (0.026) | 7.43 (0.16) | 1.348 (0.027) |

Genetic associations with HbA1c are similar across strata, with a 1.21-fold difference for the lowest versus highest strata. For SBP, there was a 1.56-fold difference, for eGFR, there was a 2.04-fold difference, and for BMI, there was a 3.66-fold difference. The constant genetic effect assumption may be a reasonable approximation for HbA1c, but it is not reasonable for BMI. For SBP and BMI, genetic associations increased monotonically across the distribution of the trait. For HbA1c and eGFR, genetic associations were similar in the lower deciles, but increased (for HbA1c) or decreased (for eGFR) in the higher deciles.

When dividing into 100 strata, evidence for a protective effect of increased BMI on all-cause mortality at very low levels of BMI as reported in the original publication [10] was less strong using the doubly-ranked method compared to the residual method (Figure 5), although the estimate in the lowest percentile stratum (lowest 1%) was in the protective direction for both methods: OR per 1 standard deviation (4.8 kg/m^2^) higher genetically-predicted BMI from the residual method 0.74 (95% CI 0.57, 0.97) versus from the doubly-ranked method 0.47 (95% CI 0.23, 0.95). However, in other low BMI strata, evidence for a protective effect of increased BMI attenuated in the doubly-ranked method.

**Figure 5:**
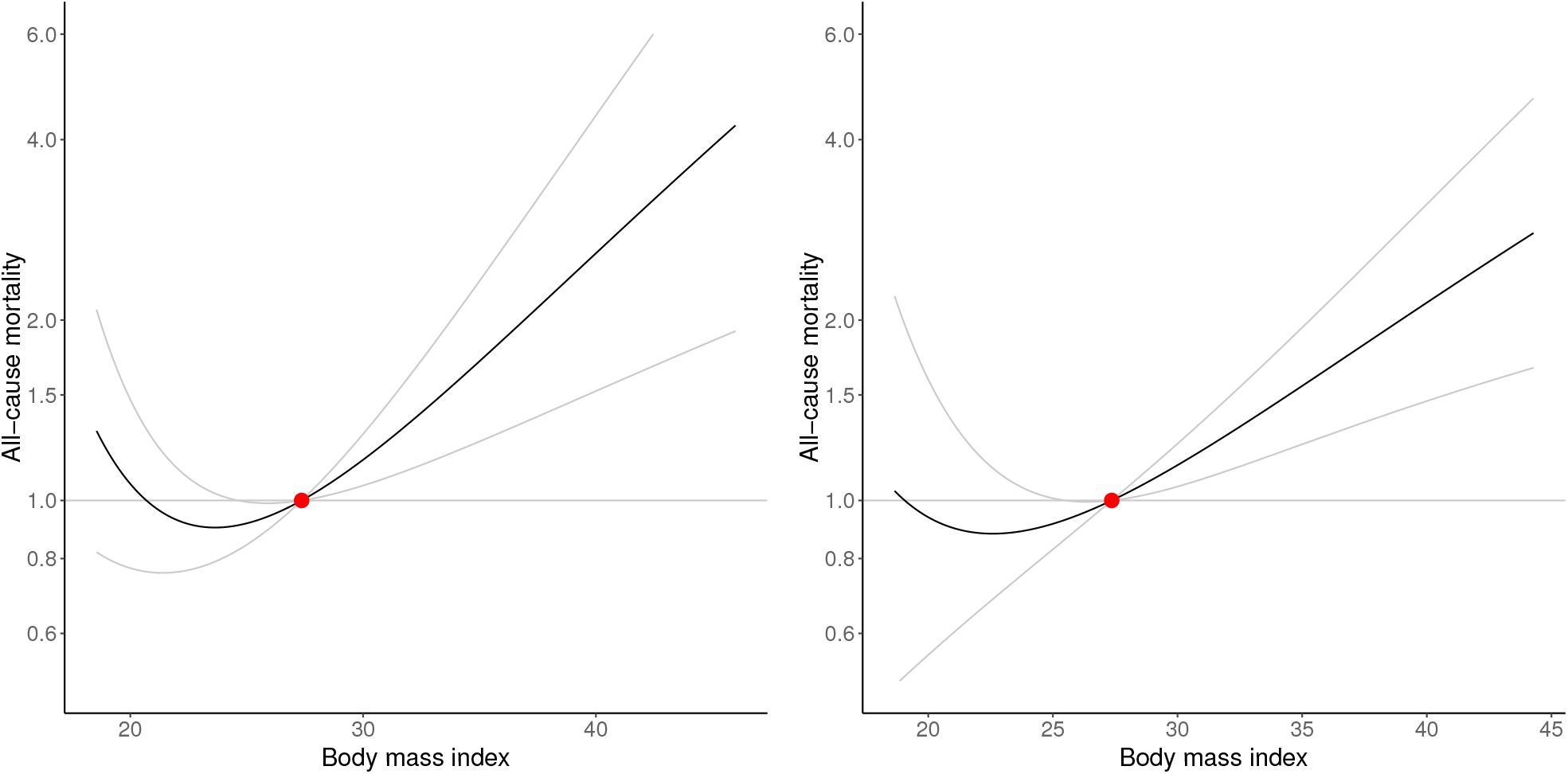
Estimated localized average causal effect curve of body mass index (BMI, kg/m^2^) on odds of all-cause mortality from non-linear Mendelian randomization using fractional polynomial smoothing taking estimates from: (left) the residual method, and (right) the doubly-ranked method. Grey lines represent the 95% confidence interval for the localized average causal effect curve.

## Discussion

In this manuscript, we have investigated differences between non-linear Mendelian randomization estimates for the effect of vitamin D on all-cause mortality using the residual and doubly-ranked methods. While the residual method suggests a potential protective effect of vitamin D at low levels, this finding is not corroborated by the doubly-ranked method. The residual method assumes that the effect of the genetic instrument on the exposure is constant for all individuals in the population. This seems to be a plausible assumption in theory, but in practice, the doubly-ranked method suggests that the assumption does not hold in this case. Further, when log-transforming the exposure so that the genetic effect on the exposure is closer to homogeneous across the population, estimates indicating a protective effect of vitamin D are substantially attenuated even in the residual method.

We conclude that the evidence for a protective effect of vitamin D in low vitamin D strata from non-linear Mendelian randomization is suspect. It is possible that some of the attenuation in the doubly-ranked method is because the lowest stratum formed by that method contains a wider group of individuals, some of whom do not have vitamin D deficiency, whereas the lowest stratum formed by the residual method contains a narrower group consisting more uniformly of deficient individuals. However, evidence for a protective effect of vitamin D supplementation attenuated substantially when log-transforming the exposure even using the residual method, meaning that this explanation is unlikely to fully explain the discrepancy between estimates. Additionally, there was evidence of instrument invalidity within strata formed by the residual method. This was not evident either for the doubly-ranked method or the residual method when log-transforming the exposure.

Although the doubly-ranked method outperformed the residual method on average in the simulation study scenarios, it too suffered from bias when the genetic effect on the exposure was not constant. Our simulations showed that the direction of bias for both methods varied depending on the factor underlying the variability of the genetic effects, and hence the direction of bias is unpredictable in practice, as this factor cannot be known. Additionally, while associations between the genetic instrument and confounder were lower for the doubly-ranked method on average, they were not uniformly lower in all datasets. This underscores the importance of the principle of triangulation [17]; results are more reliable when they are evident from multiple approaches. It also suggests that, rather than relying on a statistical method to solve the problem of genetic effect heterogeneity, approaches such as transforming the exposure should be considered to reduce the degree of heterogeneity. For most of the exposures considered in this paper, genetic associations were stronger when levels of the exposure were greater, suggesting that a log transformation or a square-root transformation may be worth considering in many cases.

In hindsight, there were two erroneous assumptions in the published Sofianopoulou analysis. The first was the assumption that the genetic effect on the exposure is constant for all individuals in the study population. While this assumption may approximately hold for some exposures, for vitamin D this assumption was sharply violated, with large differences between the estimated genetic associations across the distribution of 25(OH)D. The ratio of the genetic association estimates with the exposure in the lowest versus highest stratum was 5.00; and the ratio in the 3rd versus 8th stratum was 2.21. In contrast, corresponding ratios for HbA1c were 1.21 and 1.12, indicating that this assumption is far more reasonable for HbA1c. A brief survey of exposures indicated that violation of the constant genetic effect assumption is not isolated to 25(OH)D, although the violation was most severe for 25(OH)D out of the exposures considered. In particular, it was also strongly violated for BMI. The second erroneous assumption was that genetic associations with the exposure in strata defined by the residual method are a reliable guide of the validity of the constant genetic effect assumption, which appears not to be the case in theory [6] or in practice.

While I appreciate that science is an incremental and self-correcting discipline, I regret that one of the primary conclusion from the previous investigation into the potential effect of vitamin D supplementation is likely to be misleading, as it was based on an assumption that turned out to be violated. Although we have only presented findings for all-cause mortality here, similar discrepancies in estimates are observed for other outcomes considered in the Sofianopoulou paper. Although it would be a fallacy to claim that our updated investigation proves a null effect of vitamin D, evidence supporting a causal effect in the lowest strata is inconsistent, and only evident in the analysis that is most vulnerable to bias.

For future analyses, we recommend that non-linear Mendelian randomization investigations use the doubly-ranked method to check the constant genetic effect assumption, and transform the exposure if appropriate to reduce heterogeneity in the genetic effect on the exposure. Additionally, despite some practical advantages of the residual method, if there is evidence of heterogeneity in the genetic effect on the exposure, analysts should consider using the doubly-ranked method as the primary analysis method for non-linear Mendelian randomization.

## Data Availability

All UK Biobank data are available by application to bona fide researchers at https://www.ukbiobank.ac.uk/.

## Acknowledgements

The author would like to thank Brent Richards, George Davey Smith, Adam Butterworth, and Angela Wood for comments on an earlier draft of this work. This research has been conducted using the UK Biobank Resource under Application Number 11833. Stephen Burgess is supported by the United Kingdom Research and Innovation Medical Research Council (MC UU 00002/7) and the National Institute for Health Research Cambridge Biomedical Research Centre (BRC-1215-20014). The views expressed are those of the author and not necessarily those of the National Institute for Health Research or the Department of Health and Social Care.

## Supplementary Material

We conducted a more systematic simulation study to investigate the degree of potential bias in the residual versus doubly-ranked method. We simulated data according to the same model as in Scenario 1 of Simulation study 2 from the main text, except varying the strength of the instrument and the variability of its effects on the exposure:

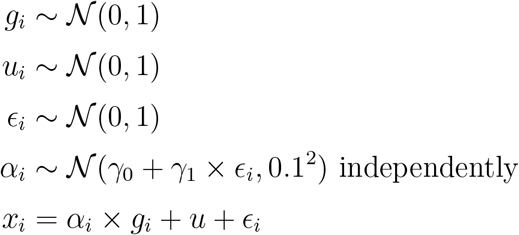

where *γ*_0_ represents the average strength of the instrument, and *γ*_1_ represents its variability. In scenarios 1, 2, and 3, we set *γ*_0_ = +0.2; in scenarios 4, 5, and 6, we set *γ*_0_ = +0.3; and in scenarios 7, 8, and 9, we set *γ*_0_ = +0.2. In scenarios 1, 4, and 7, we set *γ*_1_ = +0.05; in scenarios 2, 5, and 8, we set *γ*_1_ = +0.1; and in scenarios 3, 6, and 9, we set *γ*_1_ = +0.15. We generated 1000 datasets of 10 000 individuals for each scenario. We estimated the correlation between the genetic instrument *G* and the confounder *U* in each of 10 strata defined using the residual and doubly-ranked methods.

Results are shown in Supplementary Table A1. We see that the mean correlations in scenarios 1, 4, and 7 were similar for both methods. Similarly the mean correlations were similar in scenarios 2, 5, and 8; and scenarios 3, 6, and 9. In contrast, mean correlations were progressively higher across each set of three scenarios (1, 2, and 3; 4, 5, and 6; 7, 8, and 9) as the instrument variability was increased. This indicates that bias in Mendelian randomization estimates is dependent on the variability of the genetic effects on the exposure, not their strength.

In each scenario, the mean correlation was two times stronger for the residual method compared with the doubly-robust method, indicating that bias due to variability in the genetic effect on the exposure will be greater on average for the residual method. However, while this result held on average, in some simulated datasets correlations were stronger in the doubly-ranked method.

**Supplementary Table A1:** Mean correlations between the genetic instrument and confounder within strata in the additional simulation study for the residual and doubly-ranked methods.

| Residual method |  |  |  |  |  |  |  |  |  |
| --- | --- | --- | --- | --- | --- | --- | --- | --- | --- |
| Stratum | Scenario 1 | 2 | 3 | 4 | 5 | 6 | 7 | 8 | 9 |
| 1 | 0.065 | 0.126 | 0.184 | 0.064 | 0.126 | 0.184 | 0.064 | 0.126 | 0.183 |
| 2 | 0.050 | 0.098 | 0.146 | 0.050 | 0.098 | 0.147 | 0.050 | 0.098 | 0.146 |
| 3 | 0.032 | 0.064 | 0.097 | 0.032 | 0.065 | 0.097 | 0.033 | 0.065 | 0.097 |
| 4 | 0.019 | 0.037 | 0.056 | 0.019 | 0.037 | 0.056 | 0.019 | 0.038 | 0.056 |
| 5 | 0.006 | 0.012 | 0.018 | 0.006 | 0.012 | 0.019 | 0.006 | 0.012 | 0.019 |
| 6 | -0.006 | -0.013 | -0.018 | -0.006 | -0.012 | -0.018 | -0.006 | -0.012 | -0.019 |
| 7 | -0.019 | -0.037 | -0.056 | -0.019 | -0.037 | -0.056 | -0.019 | -0.037 | -0.056 |
| 8 | -0.032 | -0.065 | -0.097 | -0.033 | -0.064 | -0.097 | -0.032 | -0.064 | -0.098 |
| 9 | -0.050 | -0.099 | -0.146 | -0.049 | -0.098 | -0.146 | -0.050 | -0.098 | -0.146 |
| 10 | -0.064 | -0.125 | -0.183 | -0.065 | -0.126 | -0.183 | -0.065 | -0.125 | -0.184 |

| Doubly-ranked method |  |  |  |  |  |  |  |  |  |
| --- | --- | --- | --- | --- | --- | --- | --- | --- | --- |
| Stratum | Scenario 1 | 2 | 3 | 4 | 5 | 6 | 7 | 8 | 9 |
| 1 | 0.033 | 0.064 | 0.096 | 0.032 | 0.064 | 0.096 | 0.032 | 0.065 | 0.096 |
| 2 | 0.022 | 0.044 | 0.066 | 0.022 | 0.044 | 0.066 | 0.022 | 0.045 | 0.066 |
| 3 | 0.015 | 0.029 | 0.043 | 0.015 | 0.030 | 0.044 | 0.015 | 0.030 | 0.044 |
| 4 | 0.009 | 0.017 | 0.025 | 0.008 | 0.017 | 0.026 | 0.009 | 0.017 | 0.026 |
| 5 | 0.003 | 0.005 | 0.008 | 0.002 | 0.006 | 0.009 | 0.003 | 0.006 | 0.008 |
| 6 | -0.003 | -0.006 | -0.008 | -0.003 | -0.006 | -0.008 | -0.003 | -0.006 | -0.009 |
| 7 | -0.009 | -0.017 | -0.025 | -0.009 | -0.017 | -0.025 | -0.009 | -0.017 | -0.025 |
| 8 | -0.015 | -0.030 | -0.044 | -0.015 | -0.029 | -0.044 | -0.015 | -0.029 | -0.044 |
| 9 | -0.022 | -0.044 | -0.066 | -0.022 | -0.044 | -0.066 | -0.022 | -0.044 | -0.066 |
| 10 | -0.032 | -0.064 | -0.096 | -0.033 | -0.065 | -0.096 | -0.033 | -0.064 | -0.097 |

**Supplementary Figure A1:**
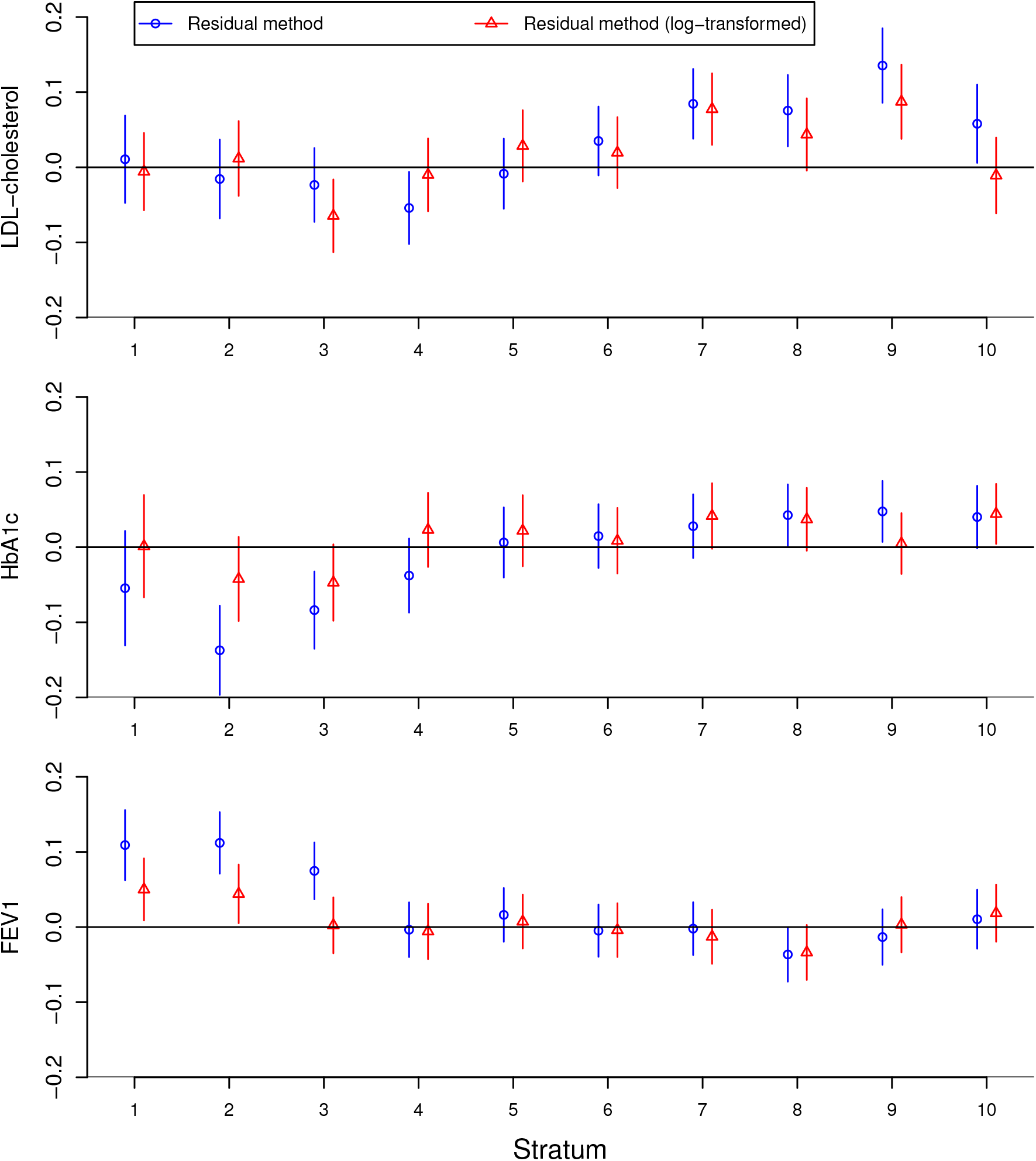
Genetic associations with covariates in strata defined by the residual method for untransformed 25(OH)D (blue circles) and log-transformed 25(OH)D (red triangles). Estimates represent associations with the covariate in standard deviation units per unit change in the genetic instrument, corresponding to a standard deviation change in 25(OH)D concentrations. Error bars are 95% confidence intervals.

## Notes

### Competing Interest Statement

The authors have declared no competing interest.

### Author Declarations

This paper uses openly available human data from UK Biobank available by application to bona fide researchers at https://www.ukbiobank.ac.uk/.

### Summary of Updates

The list of individuals included in the acknowledgement has been updated.

## References

[1] Davey Smith G, Ebrahim S. ‘Mendelian randomization’: can genetic epidemiology contribute to understanding environmental determinants of disease? International Journal of Epidemiology 2003; 32(1):1–22, doi:10.1093/ije/dyg070.

[2] Hernán M, Robins J. Instruments for causal inference: an epidemiologist’s dream? Epidemiology 2006; 17(4):360–372, doi:10.1097/01.ede.0000222409.00878.37.

[3] Burgess S, Davies NM, Thompson SG, EPIC-InterAct Consortium. Instrumental variable analysis with a nonlinear exposure–outcome relationship. Epidemiology 2014; 25(6):877– 885, doi:10.1097/ede.0000000000000161.

[4] Small DS. Commentary: Interpretation and sensitivity analysis for the localized average causal effect curve. Epidemiology 2014; 25(6):886–888, doi:10.1097/ede.0000000000000187.

[5] Cole SR, Platt RW, Schisterman EF, Chu H, Westreich D, Richardson D, Poole C. Illustrating bias due to conditioning on a collider. International Journal of Epidemiology 2010; 39(2):417–420, doi:10.1093/ije/dyp334.

[6] Tian H, Mason AM, Liu C, Burgess S. Relaxing parametric assumptions for non-linear Mendelian randomization using a doubly-ranked stratification method. bioRxiv 2022; 2022.06.28.497930, doi:10.1101/2022.06.28.497930.

[7] Staley JR, Burgess S. Semiparametric methods for estimation of a nonlinear exposure-outcome relationship using instrumental variables with application to Mendelian randomization. Genetic Epidemiology 2017; 41(4):341–352, doi:10.1002/gepi.22041.

[8] Burgess S, Malik R, Liu B, Mason AM, Georgakis MK, Dichgans M, Gill D. Dose-response relationship between genetically proxied average blood glucose levels and incident coronary heart disease in individuals without diabetes mellitus. Diabetologia 2021; 64(4):845–849, doi:10.1007/s00125-020-05377-0.

[9] Malik R, Georgakis MK, Vujkovic M, Damrauer SM, Elliott P, Karhunen V, Giontella A, Fava C, Hellwege JN, Shuey MM, et al. Relationship between blood pressure and incident cardiovascular disease: linear and nonlinear Mendelian randomization analyses. Hypertension 2021; 77(6):2004–2013, doi:10.1161/hypertensionaha.120.16534.

[10] Sun YQ, Burgess S, Staley JR, Wood AM, Bell S, Kaptoge SK, Guo Q, Bolton TR, Mason AM, Butterworth AS, et al. Body mass index and all cause mortality in HUNT and UK Biobank studies: linear and non-linear mendelian randomisation analyses. British Medical Journal 2019; 364:l1042.

[11] Luo S, Au Yeung SL, Schooling CM. Assessing the linear and non-linear association of HbA1c with cardiovascular disease: a Mendelian randomisation study. Diabetologia 2021; 64(11):2502–2510, doi:10.1007/s00125-021-05537-w.

[12] Arvanitis M, Qi G, Bhatt DL, Post WS, Chatterjee N, Battle A, McEvoy JW. Linear and nonlinear Mendelian randomization analyses of the association between diastolic blood pressure and cardiovascular events: the J-curve revisited. Circulation 2021; 143(9):895– 906, doi:10.1161/circulationaha.120.049819.

[13] Zhou A, Selvanayagam JB, Hyppönen E. Non-linear Mendelian randomization analyses support a role for vitamin D deficiency in cardiovascular disease risk. European Heart Journal 2022; 43(18):1731–1739.

[14] Biddinger KJ, Emdin CA, Haas ME, Wang M, Hindy G, Ellinor PT, Kathiresan S, Khera AV, Aragam KG. Association of habitual alcohol intake with risk of cardiovascular disease. JAMA Network Open 2022; 5(3):e223.849, doi:10.1001/jamanetworkopen.2022.3849.

[15] Gaziano L, et al. Mild-to-moderate kidney dysfunction and cardiovascular disease: observational and Mendelian randomisation analyses. Circulation 2022; In press.

[16] Sofianopoulou E, et al. Estimating dose–response relationships for vitamin D with coronary heart disease, stroke, and all-cause mortality: observational and Mendelian randomisation analyses. The Lancet Diabetes & Endocrinology 2021; 9(12):837–846, doi: 10.1016/s2213-8587(21)00263-1.

[17] Lawlor DA, Tilling K, Davey Smith G. Triangulation in aetiological epidemiology. International Journal of Epidemiology 2016; 45(6):1866–1886, doi:10.1093/ije/dyw314.

